# IgG antibody responses against antigenic salivary peptides from subgenus *Nyssorhynchus* and *Anopheles* vectors in Grand’Anse, Haiti

**DOI:** 10.64898/2026.04.01.26350006

**Authors:** Alyssa R. Schwinn, Will Eaton, Sara Harris, Vena Joseph, Alexandre Existe, Jacques Boncy, Eric Rogier, Michelle A. Chang, Daniel Impoinvil, Ruth A. Ashton, Thomas Druetz, Thomas P. Eisele, Berlin Londono-Renteria

## Abstract

*Anopheles albimanus* (*Nyssorhynchus*) is featured as the main malaria vector on Hispaniola. However, five other *Anopheles* species have been reported circulating in the area; four of them belonging to the subgenus *Anopheles* (*An. crucians, An. grabhamii, An. pseudopunctipennis*, and *An. vestitipennis*) and another one to the *Nyssorhynchus* subgenus (*An. argyritarsis*). Previous studies on mosquitoes in the genus *Anopheles* have identified and characterized peptides from immunogenic salivary proteins, with several of these peptides being unique to the *Nyssorhynchus* and *Anopheles* subgenera. This underscores their potential use as biomarkers for differentiating exposure to *Anopheles* mosquitoes in both the Old World and New World. Since both *Nyssorhynchus* and *Anopheles* subgenera have been reported in Haiti, a series of ELISAs were conducted to quantify IgG antibody titers against three published antigenic anopheline salivary peptides (gSG6-P1, Peroxi-P3, and Apy-2) in 348 participants registered in Haiti’s multi-partner/multidisciplinary Malaria Zero Program. This study aimed to evaluate the intensity of human-vector contact with *Anopheles* from both subgenera in Grand’ Anse, Haiti. In addition, the study measured antibodies against a panel of *Plasmodium falciparum* antigens to determine any association between anti-parasite and anti-peptide antibodies. Significantly elevated IgG responses to Peroxi-P3 in comparison to Apy2 and gSG6-P1 in the total study population (p < 0.001) were observed. Additionally, immune responses to Peroxi-P3 and gSG6-P1 differed significantly between ≤18-year-olds and >18-year-olds (p = 0.004 and p = 0.002), whereas no sex-based differences were observed for any peptide. Correlation analyses observed a greater number of significant positive associations in immune response between gSG6-P1 and *Plasmodium* antigens than any other salivary peptide, an occurrence which was more pronounced in ≤18-year-olds than >18-year-olds. A marked reduction in IgG responses to Apy2 and Peroxi-P3, but not gSG6-P1, among participants who kept a single household animal species compared with those who owned two or more species or those who did not have household animals was also demonstrated. Spatial analysis revealed heterogenous geographic overlap of high antibody responses among Peroxi-P3, Apy2, and gSG6-P1, alongside geographically overlapping clusters of low antibody responses to Peroxi-P3 and Apy2. These results provide additional data on the utility of anopheline salivary peptides to characterize human-vector-parasite exposure dynamics in low-transmission areas, such as Haiti.

## Introduction

Haiti continues to strive for malaria elimination with a current target date of 2025, focusing on a multipronged strategy that includes vector control measures, geographically targeted interventions, development of community-level case management, and enhancing malaria surveillance systems [1]. Several partners, including the Malaria Zero project (2016 to 2020), have contributed strategic, technical, and financial support to the strategy [1–3]. However, despite decades of dedicated efforts to eliminate malaria transmission in Haiti, it remains one of the country’s important, preventable causes of morbidity and mortality, particularly among women and children [4,5]. The complex political environment, funding instability, and frequent humanitarian crises complicate surveillance practices, making it difficult to establish an epidemiological profile for malaria transmission in the country [6].

*P. falciparum* is the primary species responsible for nearly all the uncomplicated and severe cases in Haiti, and there is thought to be limited *P. vivax* transmission, while cases of *P. malariae* infection are sporadically detected originating from the country [7–11]. *Plasmodium* parasites are transmitted through the bite of an infected female *Anopheles* spp. mosquito while taking a blood meal [12,13]. During the blood feeding process, the mosquito deposits saliva into the host to aid in the successful acquisition of blood [14]. Mosquito saliva contains hundreds of proteins, including many that possess pharmacologically active properties capable of counteracting host hemostasis through anticoagulation, vasodilation, and immunomodulation [14]. Several of these salivary proteins are immunogenic and induce antibody production closely related to the intensity of mosquito bites [14]. Therefore, previous research has used the level of such antibodies as a means of measuring mosquito exposure and risk of infection in human populations [15,16].

*Anopheles (Nyssorhynchus) albimanus* is the major malaria vector and most abundant anopheline species in Haiti [17–20]. Yet, members of the *Anopheles* subgenus, including *An. crucians, An. grabhamii, An. pseudopunctipennis*, and *An. vestitipennis* have been reported to occur in the country [19,20]. The full extent of these species as malaria vectors in Haiti is still uncertain, though mosquitoes such as *An. crucians*, *An. pseudopunctipennis*, and *An. vestitipennis* have been implicated as vectors in other regions of the western hemisphere [21–24]. For instance, *An. crucians* has been identified as a malaria vector since *P. falciparum* sporozoites have been naturally detected in its salivary glands before the elimination of malaria in the US [21,25]. However, the feeding and resting behaviors of these lesser-known species in Latin America and the Caribbean are poorly documented, though several, including *An. vestitipennis*, *An. pseudopunctipennis*, and *An. grabhamii*, are known to bite humans [24,26–28]. In contrast, *An. albimanus* has an extensively evidenced zoophilic preference, particularly for cattle, though it will opportunistically feed on humans [6,26,29–31].

Even if *Anopheles* species other than *An. albimanus* have a limited role in malaria transmission, they still bite humans along with culicine mosquitoes, and prior studies have shown that mosquito saliva itself, independent of pathogen presence, can modulate host immune responses lasting days to weeks, potentially influencing the outcome of malaria infection upon subsequent exposure to infective bites [32,33]. Although evidence on the effects of uninfected anopheline exposure on subsequent malaria outcomes is varied, and little is known about these dynamics among New World anopheline vectors, it is well established that exposure to uninfected culicines enhances viral pathogenesis in murine models [32–34]. An earlier study on *Anopheles* reported that mice pre-sensitized to the saliva of uninfected *Anopheles stephensi* exhibited elevated parasitemia and, in a susceptible murine model, an increased progression to cerebral malaria and mortality [32]. In contrast, a separate study found that mice bitten by uninfected mosquitoes before infection mounted a more robust T-helper 1 (Th1) immune response compared to naive mice, resulting in reduced liver and blood-stage parasite burdens [33]. Yet, host selection among New World anophelines is not well-defined and can be influenced by various biological and environmental factors, including altitude, infection status, pregnancy, human population density, and ecological alteration, highlighting the importance of evaluating local human-vector contact among primary and secondary vectors in malaria endemic regions [35–38].

In Latin America*, Anopheles darlingi* and *Anopheles albimanus* are among the most important malaria vectors, both belonging to the *Nyssorhynchus* subgenus [39]. In previous studies using whole salivary gland extract (SGE) from these species, participants with active malaria infection were found to have significantly higher IgG antibodies against the salivary proteins compared to healthy individuals [40,41]. Previous research in these species has identified the most immunogenic proteins in SGE and designed peptides from their immunogenic regions to optimize serological detection of antibodies against specific proteins in anopheline saliva, which is becoming increasingly important in low-transmission settings [40,41]. Two peptides, one from each species, are of particular interest for their ability to distinguish malaria-infected and uninfected populations in Colombia and their potential to serve as biomarkers for mosquito exposure in naïve populations [40,41]. These include the AnAlbPeroxi-P3 peptide (referred to as Peroxi-P3), derived from an *An. albimanus* salivary peroxidase, and the AnDarApy-2 peptide (referred to as Apy2), derived from a salivary apyrase belonging to *An. darlingi* [40,41]. Only one peptide has been characterized as a marker of exposure to mosquito bites from malaria vectors belonging to the *Anopheles* and *Cellia* subgenera, the gSG6-P1 peptide [42,43]. This peptide was designed from the amino acid sequence of salivary gland protein 6 (SG6) a highly immunogenic salivary protein in the saliva of *Anopheles gambiae* (gSG6), the major vector of malaria in Africa [43]. The gSG6-P1 peptide has been established as a suitable marker to evaluate exposure to bites from *Anopheles* species other than those belonging to the *Nyssorhynchus* subgenus, since the *Nyssorhynchus* subgenus mosquitoes do not express the SG6 proteins [44–49]. Consequently, further investigation is warranted to validate the efficacy of the salivary peptides designed from *An. albimanus* and *An. darlingi* proteins as reliable indicators of exposure to anopheline species in which SG6 is absent.

Previous studies have suggested the use of a panel of antigens to better determine exposure to *Plasmodium* parasites [50,51]. The most common antigens used are the 11-kilodalton fragment of the merozoite surface protein 1 (MSP1-19), the apical membrane antigen 1 (AMA-1), and the glutamate-rich protein (GLURP) [50,51]. Antibodies against MSP1-19 and AMA-1 present age-specific increases in seroprevalence that are strongly correlated with entomological inoculation rates (EIR), which is considered the gold standard for determining transmission intensity [50,52]. Also, early transcribed membrane proteins (Etramp) have been found highly immunogenic and useful for determining responses against *Plasmodium*, and prior research suggests Etramp 5 antigen 1 (ETR51) as a potential marker for recent parasite exposure [50,51,53,54].

Testing a panel of salivary antigens alongside *Plasmodium* markers may better estimate mosquito bite exposure and infection risk in low-transmission settings. Therefore, samples collected during the Malaria Zero project were used to evaluate the feasibility of combining three salivary antigens with *Plasmodium* markers to describe transmission in Grand’ Anse, Haiti.

The overall goal of this study was to address the limited validation of *Nyssorhynchus* salivary peptides as exposure biomarkers for SG6-absent *Anopheles* spp. using mosquitoes from Grand’ Anse, Haiti as a case.

To address the gap, the primary aim of this study:

1. Characterized and compared human-vector contact intensity between *Anopheles* and *Nyssorhynchus* subgenera by evaluating immunoglobulin (Ig) G responses to a panel of salivary peptides: gSG6-P1, Peroxi-P3, and Apy-2
2. Examined their association with a panel of short-term and long-term *Plasmodium* antigen markers: PfMSP1-19, PfAMA-1, ETRAMP 5 Ag 1, and PfGLURP R0.

The secondary aim of this study explored the associations between mosquito exposure and demographic, socioeconomic, and spatial parameters in either of the subgenera within the study population.

## Methods

### Study Sites and Participants

The samples included in this study were collected in 2017 and 2018 as part of prior research aiming to inform epidemiological patterns of malaria transmission and risk factors for infection in Grand’Anse department, Haiti [1,2]. Participant samples were selected at random from two separate studies: one of these studies was a case-control investigation, while the other focused on conducting easy access group (EAG) surveys [1,2]. In the case–control study, cases were febrile patients at participating health centers with a positive malaria rapid diagnostic test (RDT) or serological evidence of recent or past *P. falciparum* exposure, while controls were febrile patients with a negative malaria RDT and no detectable serological markers [1]. Additionally, household locations were recorded at enrollment at participating southwest Haiti health facilities and confirmed during follow-up [1]. In the EAG study, participants were recruited across two settings, schools and health facilities [2]. Individuals were eligible if they attended one of the enrolled sites, with no symptom related criteria for participation [2]. From this study population, all RDT positive and a random selection of 30% of RDT negative individual’s household location were geolocated using GPS devices (Garmin, Olathe, Kansas, USA) [2]. Across both studies, all participants were at least six months old, with protocols in place to ensure enrollment captured a broad age range of participants [1,2]. The archived samples were accessed for research purposes on February 3, 2022, and no identifiable participant information was available to the authors during or after data collection.

Grand’Anse is in the southwestern peninsula of Haiti and has recorded the highest incidence of malaria in the nation for many years, though incidence varies greatly between localities [1]. The population of Grand’Anse is just under half a million people with most of the population located on the coast [55]. The study included sites both inland and along the coast, encompassing Anse d’Hainault, Les Irois, and Dame-Marie, Chambellan, and Moron communes. Participants selected for this study were recruited across 44 sites, including 25 schools and 19 health centers. The study sites are comprised as follows: five schools and four health centers in Anse d’Hainault, four schools and three health centers in Chambellan, eight schools and five health centers in Dame-Marie, four schools and three health centers in Les Irois, four schools and two health centers in Moron, and two additional rural health centers in Petite Rivière and Mandou.

### Sample Collection

Participants provided a finger prick capillary blood sample for rapid diagnostic testing (RDT) to test for *P. falciparum* infection, as well as an additional blood sample for preparing dried blood spots (DBS) on filter paper to allow for other serological tests. At enrollment, basic demographic information (age, location of residence, sex, history of fever within the past two weeks, vector control methods in the home, etc.) was collected by the study team using a questionnaire on a mobile data collection platform (Commcare, Dimagi, Cambridge, MA, USA). For case-control participants, household follow-up visits gathered additional information regarding occupation of the household head, livestock ownership, bed net usage, and household size were collected. For EAG participants, this information was also collected at the time of enrollment.

### Antigen Preparation

Procedures describing the identification, selection, and purification of the salivary peptides used in this study have previously been published elsewhere [40,41,43]. In brief, proteins were assessed for the presence of signal peptide to *Anopheles* using an online BLAST program. Sequence and structure analyses were performed on the top *Anopheles*-specific proteins including analyses for antigenic regions, flexible (turn) regions, hydrophobic regions, stability, charge density, and linear B-cell epitopes. Those which were specific to *Anopheles* and found to be stable, flexible, charged, highly antigenic, and demonstrated <50% hydrophobicity were used for the selection of 18-22 amino acid peptides. The peptides of interest, gSG6-P1, Apy2, and Peroxi-P3, were sent for synthesis (GenScript Biotech, Piscataway, NJ, USA) and were received in lyophilized form, dissolved in ultrapure water, and stored at -20°C until use.

### Serological Screening for Plasmodium Antigens

A detailed description of serological testing procedures against *Plasmodium* antigens is provided in earlier publications [1,56,57]. To summarize, IgG antibody levels were tested against 4 *Plasmodium falciparum* antigens (PfAMA-1, PfMSP1-19, PfETR51, and PfGLURP R0) among all selected participants. IgG detection to parasite antigens of interest were conducted using a one-step multiplex bead assay (MBA) where antigens were covalently coupled to unique bead regions. The bead mix was coated to a 96-well plate (Bio-Rad, Hercules, CA, USA) at 50µL/well and washed twice with 100µL PBST (PBS 1×, Tween 20 0.05%). After washing, beads were resuspended in 50 µL antibody detection mix and 50µL of DBS sample was added. Plates were incubated overnight on a plate shaker at room temperature and shielded from the light. The following day plates were washed three times, and the beads were resuspended in 100µL 1×PBS. After resuspension, median fluorescence intensity (MFI) was measured, corrected for background reactivity, and log-transformed for each sample.

### ELISA Testing Against Anopheline Salivary Peptides (gSG6-P1, Apy2, and Peroxi-P3)

ELISA conditions were standardized as described elsewhere [15]. Eluted dried blood samples (EDBS) were prepared by eluting 1/4-inch circular hole punch of a protein saver Whatman card into 1000 µL of PBST and incubated overnight at 4°C. UltraCruz High Binding ELISA Multiwell Microplates (Santa Cruz Biotechnology, Inc, Dallas, TX, USA) were coated with 100 µL/well of gSG6-P1, Apy2, and Peroxi-P3 peptides (2 μg/mL) diluted in 1× PBS. Plates were incubated overnight at 4°C and blocked with 100 µL of 2% skim milk solution in PBS-Tween 20 (0.05%) (blocking buffer) for 1.5 h at 37°C. Individual EDBS (100 µL/well) were added to the plate in duplicate. Plates were incubated at 4°C overnight, washed three times, then incubated 2 h at 37°C with 100 µL/well of a 1:1000 dilution of goat polyclonal anti-human IgG conjugated with horseradish peroxidase (Abcam, Cambridge, MA, USA). After three final washes, colorimetric development was carried out using tetra-methyl-benzidine (TMB) (Abcam, Cambridge, MA, USA) as a substrate and the reaction was stopped with 0.25 M sulfuric acid, and the optical density (OD) was measured at 450 nm. Each assessed microplate contained in duplicate: a positive control, a negative control, and a blank. The positive control was a pool of EDBS of people with positive *P. falciparum* diagnosis. The negative control were duplicate wells with antigen and no human sample to test for any unspecific development of color. The blank was composed of wells containing no antigen and no sample to control for background signal and allow for plate-to-plate normalization.

### Plate-to-Plate Standardization

All serological data was entered into a Microsoft Access database. After, OD normalization and plate to plate variation was performed as described elsewhere [44]. Briefly, antibody levels were expressed as the ΔOD value: ΔOD = ODx − ODb, where ODx represents the mean of individual OD in both antigen wells and ODb the mean of the blank wells. For each tested peptide, positive controls of each plate were averaged and divided by the average of the ODx of the positive control for each plate to obtain a normalization factor for each plate as previously described. Each plate normalization factor was multiplied by plate sample ΔOD to obtain normalized ΔOD that were used in statistical analyses. Assay variation of samples (inter and intra assay) tested in the study was below 20% and were only included in the analysis if serum samples demonstrated a coefficient of variation ≤ 20% between duplicates.

### Statistical Analysis

Shapiro–Wilk tests were performed on all continuous variables to determine normality. All variables were found to be significant (p < 0.05), indicating non-parametric distributions. As such, Kruskal-Wallis tests were used to evaluate differences in IgG response across three or more variables. If significant, Dunn’s test was used to perform pairwise comparisons between independent groups. Mann–Whitney tests were used to assess differences among two groups, including evaluating immune responses by sex and age group. Participants were stratified into age groups selected a priori based on expected differences in exposure patterns associated with age-related behavioral factors. Bonferroni corrections were applied when necessary to adjust for multiple comparisons. Spearman’s correlation coefficients were calculated to determine the strength of association between two variables. All statistical testing was performed using R statistical software (R Foundation for Statistical Computing, Vienna, Austria) version 4.4.1 at an α level of 0.05 [58].

### Geospatial Analysis

The objective of the geospatial analysis was to determine whether significant spatial clustering in antibody response was observed in the study region. An interpolated surface was generated for descriptive purposes only to aid in visualization of trends in spatial clustering strength. Analyses were performed in R version 4.4.2 using household GPS data from 234 participants [58]. To protect individual privacy, household locations were randomly jittered between 250 meters to 1-kilometer radius in a random direction in visualizations. Statistically significant clusters of high IgG titers (hot spots) and statistically significant clusters of low IgG titers (cold spots) to each salivary peptide (Apy2, Peroxi-P3, and gSG6-P1) were identified using the Getis-OrdGi* statistic using the ‘spdep’ R package [59]. Spatial relationships of IgG responses to each peptide among participant household locations were defined using k-nearest neighbors (kNN) approach where each observation was assigned k=15 nearest neighbors. To generate a continuous predictive surface of predicted IgG responses, values at unsampled locations were interpolated using inverse distance weighted (IDW) interpolation from the ‘gstat’ R package with inverse distance power set at 4 [60].

### Ethical Considerations

The study protocols for original sample collection and survey procedures were reviewed and approved by the National Bioethics Committee of the Haitian Ministry of Public Health and Population (1718-20; 1516-30), the Tulane University Institutional Review Board (2017-366; 795709), and the London School of Hygiene and Tropical Medicine Ethics Committee (14556; 103939) [1,2]. Consent from all participants in the case-control study was recorded electronically during recruitment and confirmed at follow-up. Parental written consent was provided using an opt-out process for school surveys conducted in the EAG study. In all other settings, parents or guardians gave written consent for participants under the age of 18 years. In some cases, mature minors (aged 16-17 and either head of household, a parent, or pregnant) were able to provide written consent directly. Any participants with a positive RDT result received treatment for malaria infection upon testing positive.

## Results

### Demographic characteristics of the study population

The main characteristics of the study population are presented in Table 1. Throughout a two-year sampling period, dried blood samples were collected from 348 participants across two studies in Grand’Anse, Haiti. A total of 179 individuals were recruited from four health centers as part of a case control study conducted in 2018 in which every participant presented fever within 2 weeks prior to recruitment. The remaining 169 individuals were recruited across two years, 2017 and 2018, as part of an EAG survey pilot. Among both studies 228 (65.5%) participants were 18 years of age or younger and 120 (34.5%) participants which were 19 years or older. A histogram depicting the age distribution of the study population is provided in Figure S1. However, representation by sex was nearly equal in the combined study population with 163 (46.8%) female and 183 (52.6%) male participants. 215 (61.8%) of the participants had experienced a fever within two weeks of enrollment compared to 129 (37.1%) who had not had a fever during this time. Conversely, 262 (75.3%) people had negative RDT tests for *Plasmodium falciparum* malaria and only 85 (24.4%) were positive. RDT test results were not available for one (0.3%) person. Most of the participants (47.4%) reported sleeping under bednets the night before being enrolled in the study, though a large percentage (29.0%) did not report this information, and a smaller proportion of people (23.6%) did not sleep under bednets. Considering the large percentage of individuals for which this information was not available, no analyses were conducted based on this variable alone. There were more participants (56.6%) from households who owned farm animals than participants who did not (43.4%). Of those who did own animals, 103 (29.6%) owned only one type of farm animal while the other 94 (27.0%) people had 2 or more types of animals among the household.

**Table 1.** Characteristics of participants in total study population and in each study. Percents are expressed as the proportion of individuals among each category relative to the study population in which they were enrolled.

|  |  | **Total Study Population**<br>**n (%)** | **EAG Study**<br>**n (%)** | **Case Control Study**<br>**n (%)** |
| --- | --- | --- | --- | --- |
| **Age** |  |  |  |  |
|  | ≤18 years old | 228 (65.5) | 135 (79.9) | 93 (52.0) |
|  | >18 years old | 120 (34.5) | 34 (20.1) | 86 (48.0) |
| **Sex** |  |  |  |  |
|  | Females | 163 (46.8) | 79 (46.7) | 84 (46.9) |
|  | Males | 183 (52.6) | 88 (52.1) | 95 (53.1) |
|  | Not available | 2 (0.6) | 2 (1.2) | 0 |
| **Fever within the previous two weeks** |  |  |  |  |
|  | No | 129 (37.1) | 129 (76.3) | 0 |
|  | Yes | 215 (61.8) | 36 (21.3) | 179 (100.0) |
|  | Not available | 4 (1.1) | 4 (2.4) | 0 |
| **RDT result** |  |  |  |  |
|  | Negative | 262 (75.3) | 133 (78.7) | 129 (72.1) |
|  | Positive | 85 (24.4) | 35 (20.7) | 50 (27.9) |
|  | Not available | 1 (0.3) | 1 (0.6) | 0 |
| **Slept under a bednet the previous night** |  |  |  |  |
|  | No | 82 (23.6) | 18 (10.6) | 64 (35.7) |
|  | Yes | 165 (47.4) | 77 (45.6) | 88 (49.2) |
|  | Not available | 101 (29.0) | 74 (43.8) | 27 (15.1) |
| **Number of bednets among the household** |  |  |  |  |
|  | 0-3 bednets | 238 (68.4) | 78 (46.1) | 160 (89.4) |
|  | >3 bednets | 36 (10.3) | 17 (10.1) | 19 (10.6) |
|  | Not available | 74 (21.3) | 74 (43.8) | 0 |
| **Animals in the household** |  |  |  |  |
|  | No animals | 151 (43.4) | 81 (47.9) | 70 (39.1) |
|  | 1 type of animal | 103 (29.6) | 47 (27.8) | 56 (31.3) |

|  | Total Study Population<br>n (%) | EAG Study<br>n (%) | Case Control Study<br>n (%) |
| --- | --- | --- | --- |
| ≥ 2 types of animals | 94 (27.0) | 41 (24.3) | 53 (29.6) |
| **Household size** |  |  |  |
| 1-5 people | 184 (52.9) | 73 (43.2) | 111 (62.0) |
| 6-10 people | 149 (42.8) | 85 (50.3) | 64 (35.8) |
| >10 people | 13 (3.7) | 9 (5.3) | 4 (2.2) |
| Not available | 2 (0.6) | 2 (1.2) | 0 |
| **Totals** | 348 (100.0) | 169 (48.6) | 179 (51.4) |

### IgG responses to Peroxi-P3 are elevated compared to Apy2 and gSG6-P1

The Kruskal–Wallis revealed significant differences in IgG responses among the three salivary antigens in the overall study population (p < 0.0001). Post-hoc Dunn’s tests among all 348 participants, showed that immune responses to Peroxi-P3 were significantly higher than those to both, Apy2 and gSG6-P1 (p = 0.0004 and p = 0.0001, respectively), whereas responses to Apy2 and gSG6-P1 did not differ significantly (p = 0.828) (Figure 1). Although the difference was not statistically significant, average IgG responses to gSG6-P1 were slightly higher than those to Apy2 in the study population (Figure 1).

**Figure 1.**
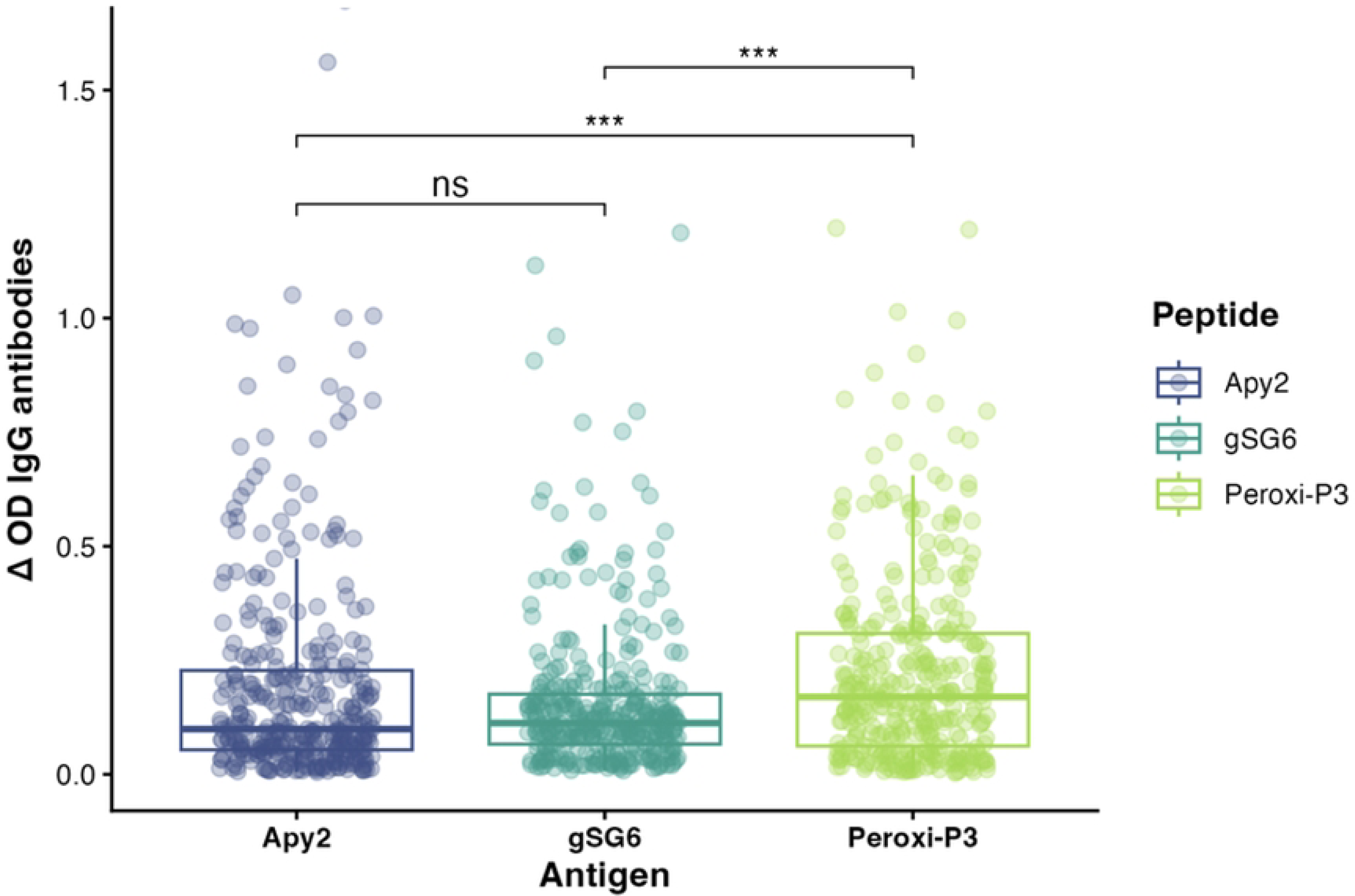
Dunn’s test comparing overall IgG antibody titers between Apy2, gSG6-P1, and Peroxi-P3 in the total study population (n=348). IgG response measured in Δ in optical density (OD) values. P-value is denoted as ***(p<0.001). NS: not significant.

### Immune responses to salivary antigens vary by age but not by sex

While IgG response was higher among male participants to every antigen, the differences were not significantly higher than the response in female participants (Figure 2A). In addition, we examined differences in IgG response to each salivary antigen among ≤18-year-old and >18-year-old study groups (Figure 2B). Similarly, immune responses to every antigen were higher ≤18-year-olds in comparison to >18-year-olds, though these differences were only significant when comparing responses to the gSG6-P1 (p = 0.002) and Peroxi-P3 (p = 0.004) peptides (Figure 2B). Finer-scale age-dependent differences were observed, with participants aged 7–17 exhibiting significantly higher responses across all antigens, whereas no differences in IgG levels were detected between the youngest and oldest participants for any antigen (Figure S2). No significant differences in vector antigen exposure were detected among participants based on household size (Figure S3).

**Figure 2.**
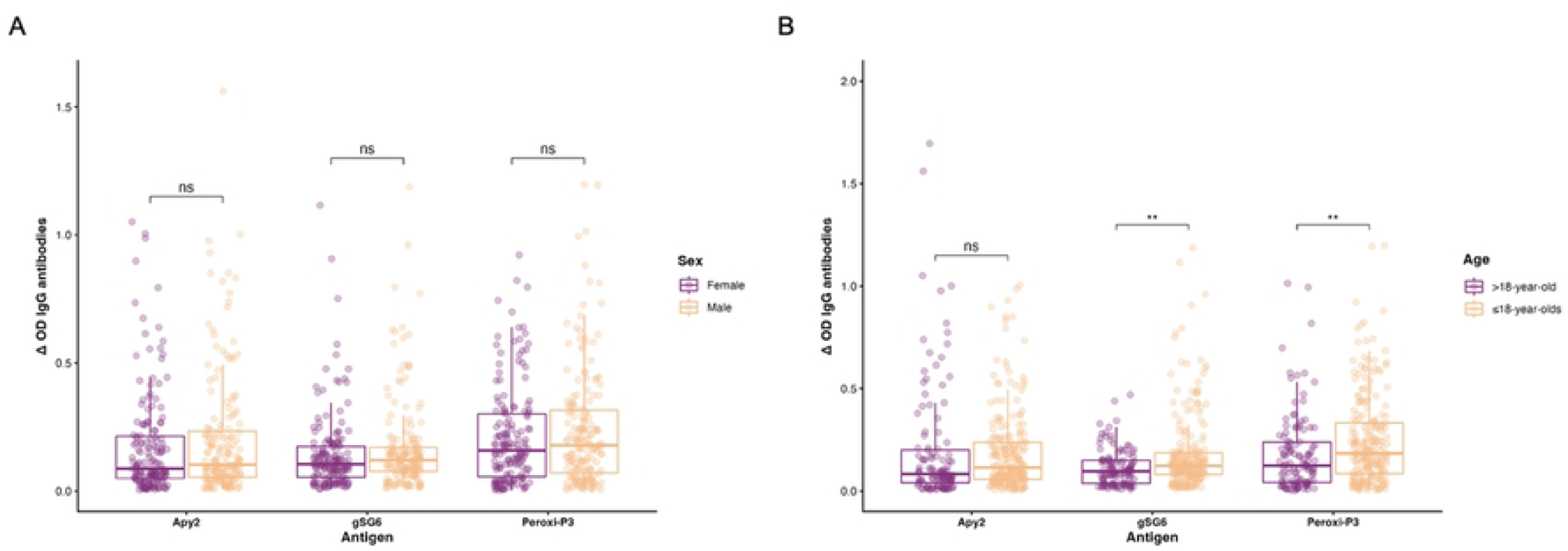
Mann-Whitney U test comparing IgG antibody titers to Apy2, gSG6-P1, and Peroxi-P3 among participants by sex (A). Mann-Whitney U test comparing IgG antibody titers to Apy2, gSG6-P1, and Peroxi-P3 among participants by age group (B). IgG response measured in Δ in optical density (OD) values. P-values are denoted as **(p<0.01). NS: not significant.

### IgG responses to gSG6-P1 are correlated with exposure to Plasmodium antigens in children

Correlation analysis of IgG responses to salivary antigens with *Plasmodium* antigens in the total study population did not find any significant associations between exposure to *Plasmodium* antigens and salivary peptides from *Anopheles* of the *Nyssorhynchus* subgenus (Apy2 and Peroxi-P3). Instead, only two significant associations occurred, revealing that both ETR51 and GLURP-R0 were positively associated with exposure to the gSG6-P1 peptide from Anophelines of the *Anopheles* or *Cellia* subgenus (ρ = 0.1781, p = 0.0009; ρ = 0.2102, p < 0.0001) (Table 2). The relationship between exposure to *Plasmodium* antigens and gSG6-P1 was more pronounced in children, where three *Plasmodium* antigens (AMA-1, ETR51, and GLURP-R0) were positively associated with exposure to the gSG6-P1 peptide (ρ = 0.238, p = 0.0003; ρ = 0.181, p = 0.0064; ρ = 0.265, p < 0.0001). IgG response to Peroxi-P3 and GLURP-R0 was also positively associated within this age group (ρ = 0.151, p = 0.0235) (Figure 3A). MSP1-19 was not significantly associated with exposure to any peptide in younger participants. In contrast, only one significant association between *Plasmodium* and *Anopheles* antigens occurred in adults. In older participants, exposure to Peroxi-P3 was negatively associated with exposure to MSP1-19 (ρ = -0.186, p = 0.0432) (Figure 3B).

**Figure 3.**
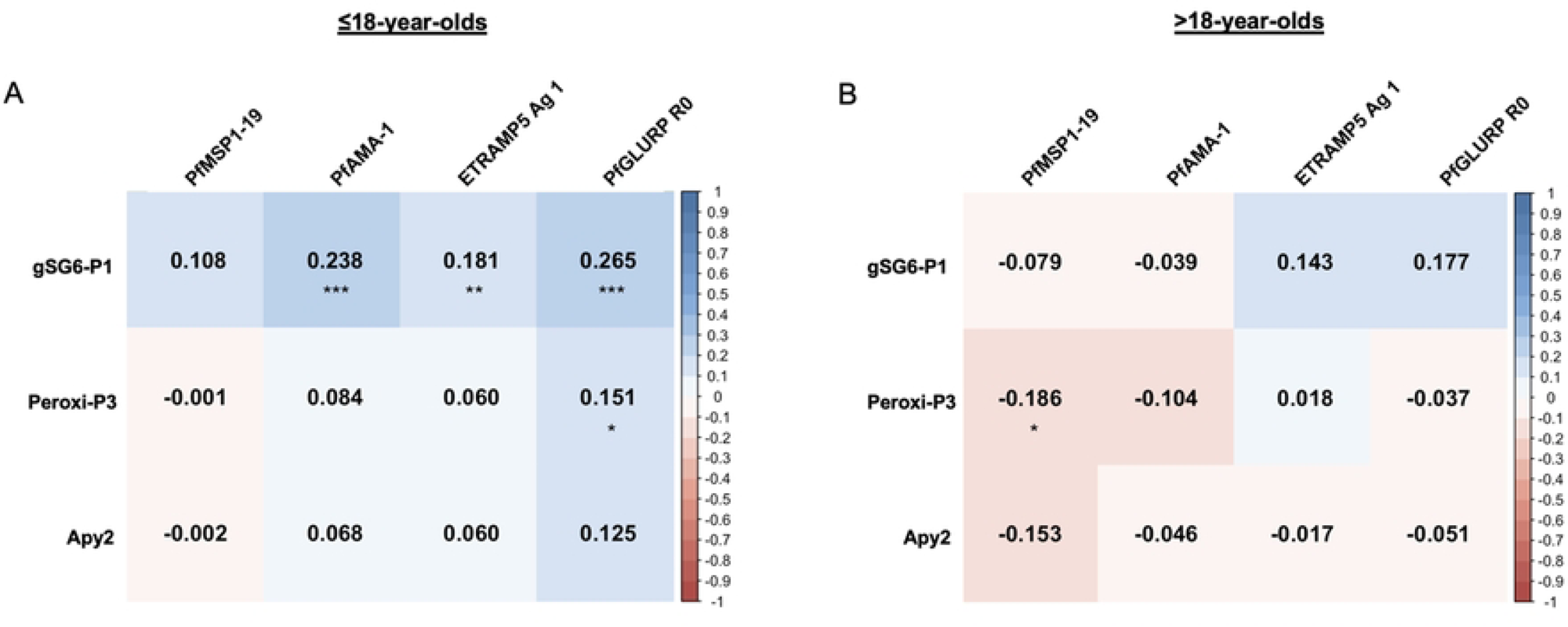
Correlogram of Spearman correlations of IgG antibody titers against *Anopheles* salivary peptides and *Plasmodium* antigens among participants 18 years or younger (n=226) (A). Correlogram of Spearman correlations of IgG antibody titers against *Anopheles* salivary peptides and *Plasmodium* antigens among participants 19 years or older (n=118) (B). Positive correlations are indicated in gradient blue according to strength of association (darker blue represents a stronger positive correlation) while negative correlations are indicated in gradient red according to strength of association (darker red represents a stronger negative correlation). IgG response measured in Δ in optical density (OD) values. P-values are denoted as *(p<0.05), **(p<0.01), ***(p<0.001). Relationships without an asterisk are not significant.

**Table 2.**
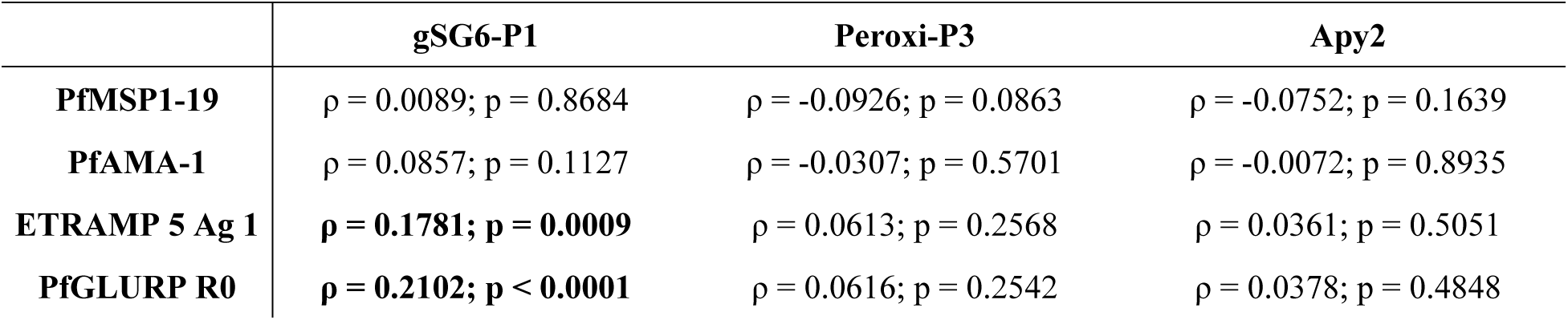
Results of all Spearman correlations of IgG antibody titers against gSG6-P1, Peroxi-P3, and Apy2 and each *Plasmodium* antigen (PfMSP1-19, PfAMA-1, ETRAMP5 Ag 1, PfGLURP R0) among all participants included in the study for which these values were available (n=344).

|  | gSG6-P1 | Peroxi-P3 | Apy2 |
| --- | --- | --- | --- |
| **PfMSP1-19** | $\rho = 0.0089$; $p = 0.8684$ | $\rho = -0.0926$; $p = 0.0863$ | $\rho = -0.0752$; $p = 0.1639$ |
| **PfAMA-1** | $\rho = 0.0857$; $p = 0.1127$ | $\rho = -0.0307$; $p = 0.5701$ | $\rho = -0.0072$; $p = 0.8935$ |
| **ETRAMP 5 Ag 1** | **$\rho = 0.1781$; $p = 0.0009$** | $\rho = 0.0613$; $p = 0.2568$ | $\rho = 0.0361$; $p = 0.5051$ |
| **PfGLURP R0** | **$\rho = 0.2102$; $p < 0.0001$** | $\rho = 0.0616$; $p = 0.2542$ | $\rho = 0.0378$; $p = 0.4848$ |

### Ownership of a single household animal species may protect against exposure to Anopheles (Nyssorhynchus) mosquitoes but not those of subgenus Anopheles

IgG responses were tested according to number of household animal species to determine if increased animal diversity within a household influenced differential distributions of exposure to each salivary antigen. When participants were organized in this manner, those who owned one species of animal had significantly lower exposure to both *Nyssorhynchus* peptides, Apy2 and Peroxi-P3, compared to those who did not have any animals (p = 0.0006 and p = 0.009, respectively). This difference was not observed between participants who did not own animals and those who owned two or more different types of animals to either peptide. No differences were found between those who owned one type of animal and those who owned two or more to these antigens. IgG responses to gSG6-P1 did not differ among any group according to number of animal species in the home (Figure 4).

**Figure 4.**
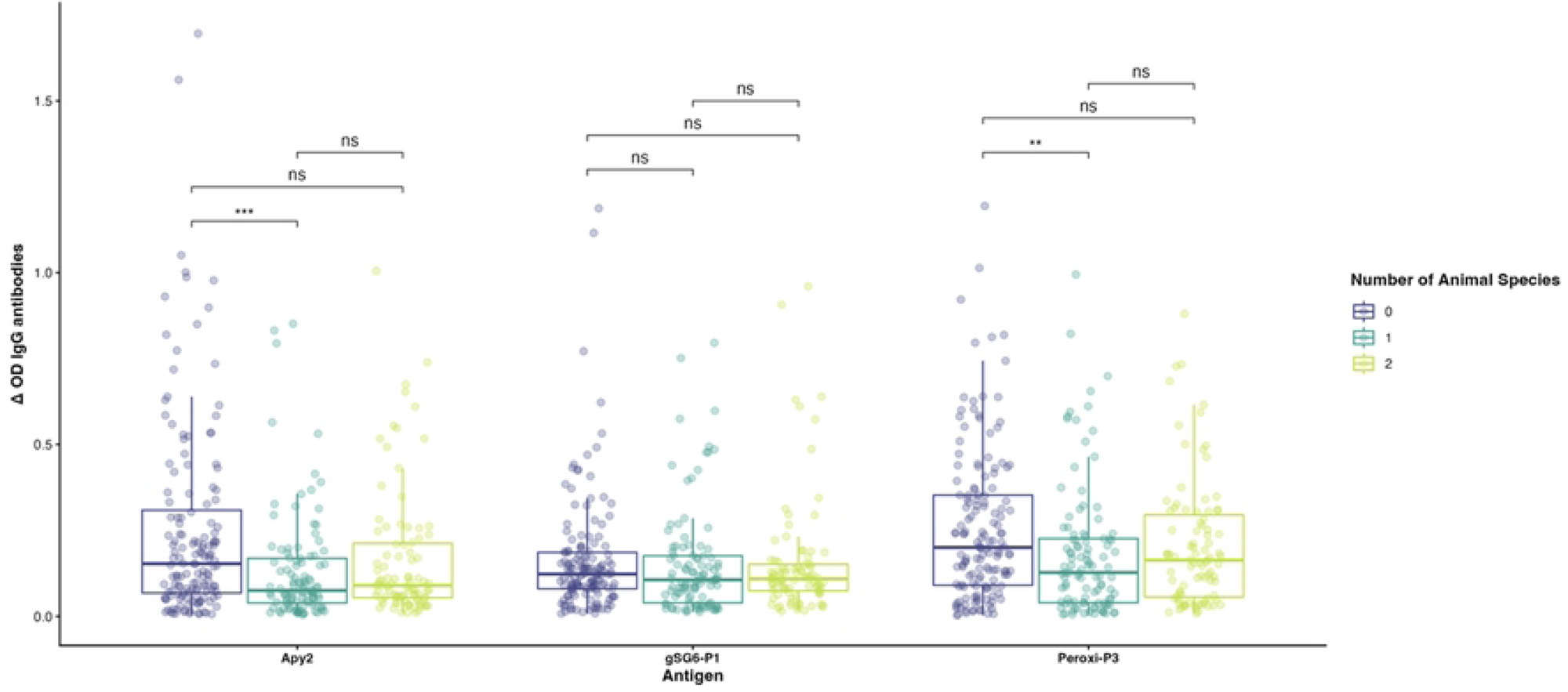
Mann-Whitney U test comparing IgG antibody titers according to number of household animal species to Apy2, gSG6-P1, and Peroxi-P3 among total study population (n=348). Group “0” (navy) is representative of households with no animals, Group “1” (teal) represents households with only a single animal species, and Group “2” (green) represents households with 2-5 animal species. IgG response measured in Δ in optical density (OD) values. P-values are denoted as **(p = 0.009) and ***(p = 0.0006). NS: not significant.

### Hotspots of *Nyssorhynchus* and *Anopheles* subgenera exposure overlap

Individuals with higher IgG antibodies against Apy2 and Peroxi-P3 salivary peptides, tended to reside along the coast of Dame Marie, Anse d’Hainault, and Les Irois communes within Grand’Anse department (Figure 5). Individuals with lower values of antibodies against Apy2 and Peroxi-P3 salivary peptides appeared more evenly distributed among all western Grand’Anse communes suggesting low-level exposures to *Nyssorhynchus* subgenera mosquitoes including *An. albimanus*, throughout much of western Grand’Anse (Figure 5, Row 1). Hotspot analysis results revealed clusters of greater exposure to *Nyssorhynchus* mosquitoes along coastal areas of Anse d’Hainault and northern Les Irois communes, as well as the inland areas of Dame Marie and Chambellan communes. Coldspots or clusters of lower exposure to *Nyssorhynchus* subgenera mosquitoes was observed along coastal areas of northern Dame Marie commune and southern coastal areas of Anse d’Hainault. Additionally, hotspots of exposure to *Anopheles* subgenera mosquitoes were observed along the coast of Les Irois and within inland areas of Chambellan commune (Figure 5, Row 2).

**Figure 5.**
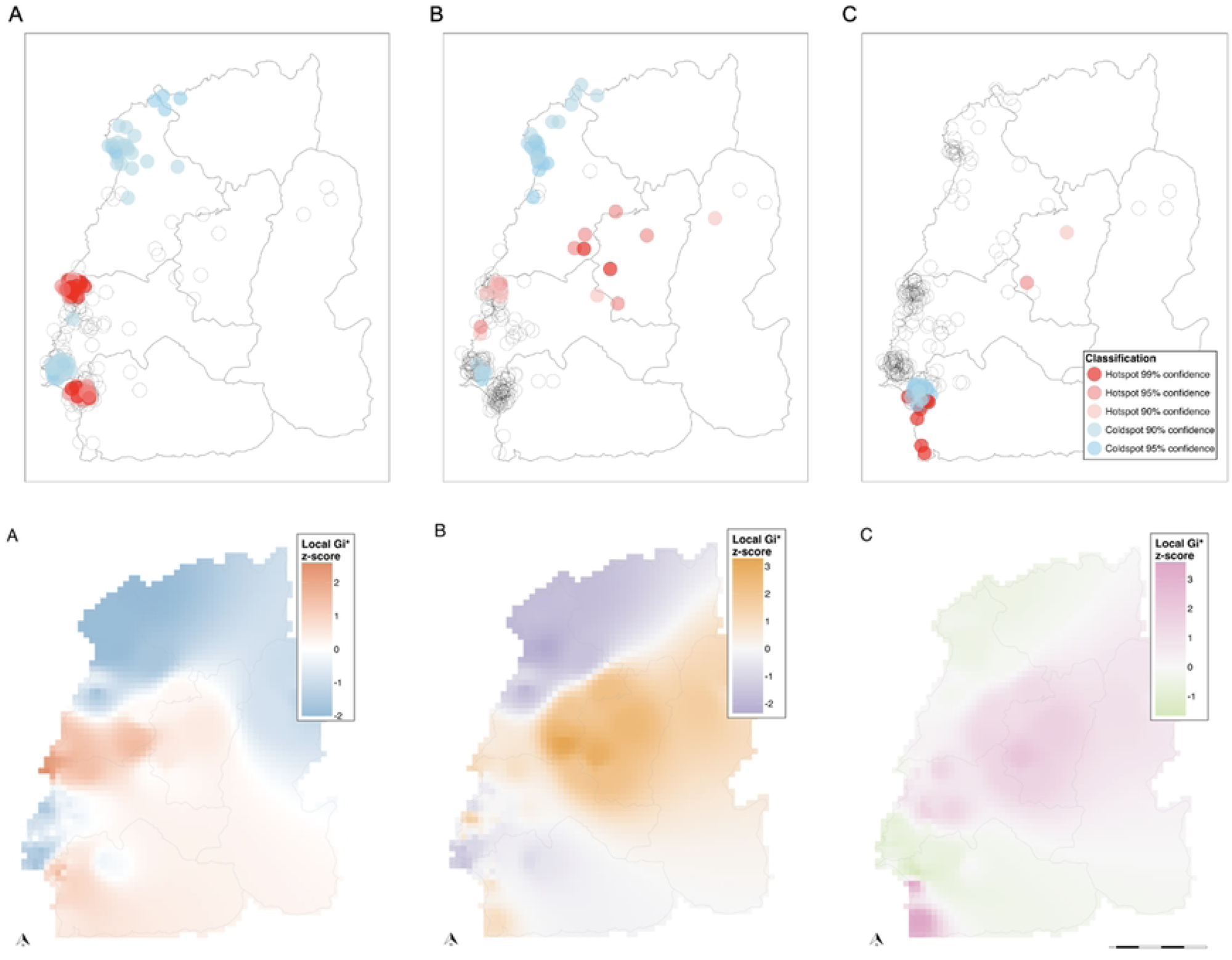
Row 1. Spatial distribution of hotspot analysis results of IgG antibodies against A) Apy2, B) Peroxi-P3, and C) gSG6-P1. Points represent classified locations identified through Getis-Ord Gi* hotspot analysis, with confidence intervals denoting hot or cold spot strength. All point locations have been spatially jittered by a random distance between 250 meters and 1 kilometer in a random direction. **Row 2**. Spatial interpolation of local Getis-Ord Gi z-scores. The raster represents inverse distance weighted interpolations of point-based Gi* z-scores from hotspot analysis of IgG antibodies against A) Apy2, B) Peroxi-P3, and C) gSG6-P1 and is shown for visualization of spatial tendencies only.

## Discussion

In this study, we measured the antibody responses of residents from Grand’ Anse to immunogenic salivary peptides derived from two subgenera of the mosquito genus *Anopheles* (*Nyssorhynchus* and *Anopheles*). We aimed to examine the association of these responses with a panel of short-term and long-term *Plasmodium* antigen markers. We also assessed biological factors and spatial heterogeneity that may influence vector exposure in the population. Overall, the key findings showed that IgG responses to Peroxi-P3 were significantly higher than responses to gSG6-P1 and Apy2. Analysis of the geographic distribution of immune responses showed co-localized IgG hotspots for both Peroxi-P3 and gSG6-P1 in the Chambellan commune, and for Apy2 and gSG6-P1 in Les Irois. Additionally, age-related differences emerged, with ≤18-year-olds displaying stronger IgG responses to both Peroxi-P3 and gSG6-P1 compared to >18-year-olds, though no differences were present between sexes to any of the peptides. Correlation analysis demonstrated positive associations between IgG responses to gSG6-P1 and two *P. falciparum* antigens (ETR51 and GLURP R0), both indicators of recent parasite exposure. These relationships were more pronounced in ≤18-year-olds, who showed an additional positive association between gSG6-P1 and AMA-1. In contrast, only a single negative association appeared in >18-year-olds, between Peroxi-P3 and MSP1-19. Lastly, analysis of IgG response relative to household animal ownership suggests that keeping a single animal species may reduce exposure to *Nyssorhynchus* mosquitoes.

Recent entomological surveys have identified *An. albimanus* as the single most abundant mosquito species within Grand’Anse [18]. Though, four other anopheline mosquitoes have been previously reported within the country, only *An. albimanus* has satisfied Barnett’s criterion for implicating candidate disease vectors [17,61]. Therefore, it is not unexpected that IgG titers against Peroxi-P3, which indicates exposure to the *Nyssorhynchus* subgenus, would be significantly higher than titers against gSG6-P1, which indicates exposure to the secondary vectors of *Anopheles* and *Cellia* subgenera [40,62]. Additionally, evidence suggesting the absence of SG6 in the *Nyssorhynchus* subgenus reinforces the expectation that IgG responses against Peroxi-P3 would be higher than those against gSG6-P1 in Grand’ Anse [45–47]. However, the discovery of a stronger IgG response to Peroxi-P3 in comparison to Apy2 is more challenging to explain, as both peptides measure exposure to the *Nyssorhynchus* subgenus [40,41]. Earlier characterizations and comparisons of the anopheline sialome (salivary gland proteins) across three subgenera (*Cellia*, *Anopheles*, and *Nyssorhynchus*) detail the identification of a single salivary apyrase expressed in the salivary glands of female anopheline vectors, irrespective of subgenera [47]. In contrast, the examination of salivary peroxidases identified a single shared peroxidase in mosquitoes belonging to the *Cellia* and *Anopheles* subgenera but revealed a cluster of five peroxidase genes in those of the *Nyssorhynchus* subgenus [47]. Three of the genes in this cluster are regarded as encoding non-salivary peroxidases, though the possibility of their salivary expression has not been dismissed [47]. The remaining two genes were confirmed to encode for salivary peroxidases in *An. albimanus*, indicating the expression of an additional salivary peroxidase in this mosquito, providing a possible explanation for the elevated IgG response to Peroxi-P3 as compared to Apy2 [47]. Additionally, phylogenetic analyses of the sialomes of six major *Plasmodium* vectors (four of the *Cellia* subgenus and two of the *Nyssorhynchus* subgenus) determined that the salivary peroxidases of the *Nyssorhynchus* subgenus clustered distinctly from those in the *Cellia* subgenus [63]. Further, the Apy2 peptide used in this study was designed from the salivary gland antigen of *An. darlingi* and was reported in an earlier study to have no significant similarity to apyrase peptides of *An. albimanus*, corroborating our observations, which show higher levels of anti-Peroxi-P3 antibodies compared to the other two peptides and the absence of statistical significance between anti-Apy2 and anti-gSG6-P1 IgG antibody levels [40,47,64].

To further understand these responses, we examined variation across demographic subgroups, finding that age contributed to major differences in anti-Peroxi-P3 and anti-gSG6-P1 IgG titers, with ≤18-year-olds demonstrating higher responses than >18-year-olds to all salivary peptide markers. Considering that risk of disease severity is strongly influenced by host age and exposure intensity, it is expected that ≤18-year-olds have more dramatic responses to the *Anopheles* peptides [65–67]. It is likely that a larger proportion of ≤18-year-olds, particularly those younger than 7 years of age, have not had sufficient exposure to *Plasmodium* and *Anopheles* antigens to establish immunity [68]. This finding is consistent with a 2022 study in Haiti which reported children under 15 having increased odds of high *An. albimanus* anti-SGE IgG compared to participants older than 15 years [68]. Similar immunological observations were also reported in a study examining IgG responses to *Simulium damnosum* saliva in Ghana [69]. Together, these findings are likely not indicative of true exposure differences between the two subpopulations but rather suggest the development of immune tolerance in individuals with sustained exposure to *Anopheles* bites.

Correspondingly, the relationships between *Anopheles* and *P. falciparum* antigens differ between ≤18-year-olds and >18-year-olds in this study population. Jaramillo-Underwood et al. recently reported that when controlling for other covariates seroprevalence against three *P. falciparum* antigens (circumsporozoite surface protein [CSP], reticulocyte binding-like protein homologue 2 [Rh2.2030], and schizont egress antigen [SEA-1]) was associated with high *Anopheles* anti-SGE IgG responses among children (aged 0-15 years) in Artibonite, Haiti [68]. While our study found positive associations among three *P. falciparum* antigens (AMA-1, ETR51, GLURP-R0) and *Anopheles* anti-peptide IgG in ≤18-year-olds, the Artibonite study similarly identified positive associations between two of the *Plasmodium* antigens, SEA-1 and CSP, and IgG responses to *Anopheles* SGE, but reported a negative association between anti-SGE IgG and Rh2.2030 [68]. In this study, we observed a single inverse relationship between *Anopheles* and *P. falciparum* antigens occurring between Peroxi-P3 and MSP1-19 in adults, which aligns with a study recently performed in Ghana [67]. The results in both children and adults reflect the unique immunologic trends previously described. Specifically, ≤18-year-olds appear to have minimal or undeveloped immunity to *Plasmodium*, whereas >18-year-olds exhibit partial immunity, consistent with long-term exposure to the parasite. This partial immunity to long-term markers of blood-stage infection (MSP1-19) is linked with intense exposure to *Nyssorhynchus* mosquitoes as would be expected for this region. Interestingly, IgG responses to gSG6-P1 in ≤18-year-olds are positively associated with both short-term markers of infection (ETR51 and GLURP-R0) whereas only a single peptide derived from *Nyssorhynchus* displayed a relationship to *Plasmodium* in this age demographic. This could suggest malaria transmission occurring through secondary vectors in immunologically naïve populations as *An. albimanus* is the focus of vector control initiatives. In addition, the consideration that the >18-year-olds have some immunity to infection could mask associations in immune response to secondary vectors, though these hypotheses would require further investigation to state with confidence.

Sex and household size were found to have no statistically significant impact on exposure to *Anopheles* of either subgenus in this study population. This is corroborative of a recent study in Grand’ Anse where neither sex was observed to experience increased odds of exposure to *Plasmodium* based on serological markers of recent infection or RDT positivity [1]. Yet, another study in Haitian children found that girls had increased odds of high IgG response to *An. albimanus* SGE compared to boys of similar age [70]. The use of whole SGE in the latter study rather than an individual peptide as was performed here could lead to the discrepancy between the findings related to the role of sex in *Anopheles* exposure. Since SGE comprises numerous antigens that can be recognized by IgG antibodies, the measurements likely capture responses to additional epitopes beyond peroxidase and apyrase, which may be more sensitive indicators of sex-based IgG variation. As a result, we conclude that none of the peptides examined in this study are appropriate for detecting sex-specific exposure patterns or immunologic trends. Regarding household size, some evidence from India suggests that those from smaller households are at a reduced risk of malaria transmission, though there are few studies reporting these findings in the Americas [71,72]. Still, another study in western Kenya did not find that household size influenced malaria risk [73]. The inconsistent role of household size on malaria risk across differing geographical settings demonstrate the possibility of regional deviations in household socioeconomic factors contributing to malaria incidence, as well as the feeding and host-seeking behaviors of *Nyssorhynchus* and *Anopheles* subgenera mosquitoes. In any case, it appears household size is of little significance in influencing *Plasmodium* exposure in the southwest regions of Haiti.

Another household factor to consider as a potential risk for exposure was animal ownership. *An. albimanus* is historically regarded as an exophagic and zoophilic vector of malaria, so the observed influences in *Anopheles* exposure characteristics related to household livestock in southwest Haiti were anticipated and align with observations described by Ricciardi et al. in 1971 in which the human feeding rate of *An. albimanus* was reduced in areas of high livestock density [17,29,74]. Notably, 93% of engorged *An. albimanus* in those areas were identified as having fed on cattle [74]. However, the relative importance of large ruminants versus smaller animals in vector exposure in this population is unclear. Several other studies examining the host selection behaviors of *An. albimanus* detail comparable outcomes, reinforcing a clear preference for cattle and other large ruminants as a blood source [29,75,76]. Given the zoophilic nature of *An. albimanus* and earlier trends reported regarding livestock density, it is reasonable to find that ownership of at least one type of animal reduces exposure to *An. albimanus* bites. However, finding that ownership of more than one type of animal has no effect on exposure is somewhat harder to interpret though not inexplicable. It has been previously shown that *An. albimanus* mosquitoes violate assumptions of gonotrophic concordance and readily consume multiple bloodmeals within a single gonotrophic cycle [77]. This suggests that greater diversity in available hosts may promote multiple bloodmeal behavior and encourage flexibility in host-seeking or opportunistic feeding, reducing preference for any single host species.

While numerous statistically significant clusters of elevated IgG responses to each salivary peptide were identified throughout the study districts, geographic overlap existed among Peroxi-P3 and gSG6-P1 hotspots in Chambellan, in addition to Apy2 and gSG6-P1 in Les Irois, indicating exposure to both anopheline subgenera in these areas. Interestingly, Dismer et al., 2021 recently identified a malaria hotspot in southwest Les Irois in the same region where we observed hotspots of exposure to secondary vectors [55]. Additional clusters of increased IgG titers to Peroxi-P3 and Apy2 occurred in the shared coastal border of Dame Marie and Anse d’Hainault communes, which also align with hotspot malaria cumulative incidence rate clusters identified from 2014 and 2015 in Grand’Anse [55]. The observed spatial heterogeneity of IgG responses to salivary peptides associated with *Nyssorhynchus* mosquitoes could suggest regional differences in human behavior and adherence to vector control tools, vector ecology, or vector habitat suitability. Recent evidence suggests that despite the trending exophagic behavior of *An. albimanus*, LLIN distribution campaigns are effective at reducing malaria incidence in Grand’Anse [17,78]. This suggests that LLINs are effective against *An. albimanus*, though it is unclear how variability in LLIN use rates are associated with sub-commune level *Anopheles* subgenera exposure differences [78]. Additionally, the spatial alignment in IgG responses to salivary peptides associated with primary and secondary malaria vectors combined, as well as malaria incidence hotspots, show their potential in aiding to identify and target malaria transmission foci and allow for improved targeted vector and disease control interventions [55].

Collectively, our findings reveal distinct patterns of exposure to both primary and secondary malaria vectors in Grand’ Anse, while establishing foundational validation for using newly identified *Nyssorhynchus* salivary peptide markers to measure human exposure to New World anopheline mosquito bites. Furthermore, these results underscore the importance of targeted investigations into the role secondary vector species may have in maintaining or enhancing malaria transmission in Haiti, particularly as vector control strategies continually target *An. albimanus* as the primary focus.

## Strengths and Limitations

The primary strength of this study is the use of antigenic peptides to measure exposure to anopheline vectors of different subgenera more accurately, rather than using whole salivary gland extract. The exclusive use of antigenic peptides allows for a more accurate and targeted measure of exposure to a specific mosquito species as opposed to SGE, which may have shared antigenic targets across species in the same genus. The incorporation of both long-term and short-term *Plasmodium* markers of exposure also benefits the results produced by the study, especially when considering the heterogeneity of the study population demographics.

Despite these findings, the study has several important limitations that warrant consideration during data interpretation. A key limitation is the heterogeneity of the study populations included in this analysis. Particularly, it is important to acknowledge those participants who were originally recruited for the case-control study, presented with fever at the time of sample collection, irrespective of *Plasmodium* infection status. The immunological profiles observed in these febrile patients may not be representative of the broader study population. The geographic data presents another limitation, as household coordinate data were unavailable for all participants and disproportionately represented the case-control cohort, though some sites only represent participants from the EAG surveys, possibly inflating the effect of age in the hotspot analysis.

Regarding the case-control participants, this may introduce bias arising from systemic differences between healthcare-seeking and non-healthcare-seeking populations in terms of risk factors and exposure characteristics, especially those pertaining to geographic risk distribution and focal vector contact. Lastly, since participant recruitment occurred over two years, the potential impact of interannual variation in vector density on the results cannot be excluded or accurately quantified.

## Conclusion

As Haiti advances toward malaria elimination, significant regional variations in parasite prevalence, combined with political instability, fragile health systems, inadequate funding, natural disasters, logistical barriers, and sociobehavioral determinants, continue to impede progress toward a malaria-free nation and threaten elimination efforts in neighboring regions of the Dominican Republic. Little is understood about the entomological landscape of malaria in Haiti, especially regarding the role of secondary vectors in sustaining transmission in areas of relatively high prevalence. Presently, Haiti is implementing a comprehensive strategy to reduce malaria transmission that includes a focus on vector control measures. As efforts to reduce malaria prevalence continues, interrupting ongoing transmission will become increasingly important to achieve elimination targets and to inform effective intervention strategies. In this context, serological markers of mosquito exposure offer a complementary approach to traditional entomological surveillance, particularly in low-transmission and resource-limited settings. While prior studies in Haiti have characterized population-level exposure characteristics using serology against *Anopheles* (*Nyssorhynchus*) salivary proteins, few have examined the immune responses to *Anopheles* belonging to other subgenera [15,68,70]. Here, we evaluated mosquito exposure in Grand’ Anse by analyzing immune responses to three novel antigenic salivary peptides derived from two distinct anopheline subgenera, to improve the understanding of the human-vector-pathogen interface in Grand’ Anse, define biologic and ecologic factors associated with exposure, and offer preliminary validation for the serological application of these markers. However, further longitudinal studies are needed to elucidate the temporal dynamics and persistence of these antibody responses as it relates to seasonal transmission and vector control activities.

## Abbreviations

[*Pf*]AMA-1: [*Plasmodium falciparum*] apical membrane antigen 1
*An.*: *Anopheles*
[AnDar]Apy2: [*Anopheles darlingi*] apyrase peptide 2
CSP: circumsporozoite surface protein
[E]DBS: [eluted] dried blood samples
EAG: easy access groups
ELISA: enzyme linked immunosorbent assay
[*Pf*]ETR51: [*Plasmodium falciparum*] early transcribed membrane protein 5 antigen 1
[*Pf*]GLURP: [*Plasmodium falciparum*] glutamate rich protein
GPS: global positioning system
gSG6-P1: *Anopheles gambiae* salivary gland protein 6 peptide 1
IDW: inverse distance weighted
IgG: immunoglobulin G
kNN: k-nearest neighbors
LLIN: long-lasting insecticidal net
MBA: multiplex bead assay
MFI: median fluorescence intensity
[*Pf*]MSP1-19: [*Plasmodium falciparum*] merozoite surface protein 1 OD optical density
PBS: phosphate buffered saline
[AnAlb]Peroxi-P3: [*Anopheles albimanus*] peroxidase peptide 3
RDT: rapid diagnostic test
Rh2.2030: reticulocyte binding-like protein homologue 2
SEA-1: schizont egress antigen
SGE: salivary gland extract
TMB: tetra-methyl-benzidine

## Data Availability

Due to ethical restrictions related to participant privacy, data are not publicly available. Requests for de-identified data access may be directed to the corresponding author.

## Acknowledgements

We gratefully acknowledge the support and partnership of the Haitian health authorities, IMA World Health, the Malaria Zero Consortium, and the field collection teams. We also extend our sincere thanks to the study participants, local communities, and health facility personnel for their time and contributions. In addition, we thank the team at the Laboratoire National de Santé Publique in Port-au-Prince for their expertise, efforts, and dedication in conducting the serological analyses related to parasite exposure: Jacquelin Présumé, Ithamare Romilus, Gina Mondélus, and Tamara Elismé.

## Supporting Information

**Figure S1.** Age distribution and density curve among the total study population (n = 348). A bin width of 1 was applied, with each bar corresponding to the number of participants of that age at the time of sample collection.

**Figure S2.** Mann-Whitney U test comparing IgG antibody titers to Apy2, gSG6-P1, and Peroxi-P3 among participants among finer scale age groups. Group “0–6” included participants aged six years or younger (n = 63), Group “7–17” included participants aged 7 to 17 years (n = 151), and Group “18+” included participants aged 18 years or older (n = 134). IgG response measured in Δ in optical density (OD) values. P-values are denoted as *(p<0.05), **(p<0.01), ***(p<0.001), and ****(p<0.0001). NS: not significant.

**Figure S3.** Mann-Whitney U test comparing IgG antibody titers to Apy2, gSG6-P1, and Peroxi-P3 among participants according to household size. Group “1–4” included participants from households with one to four members (n = 114), Group “5–6” included those from households with five or six members (n = 170), and Group “8–15” included participants from households with eight to fifteen members (n = 64). IgG response measured in Δ in optical density (OD) values. NS: not significant.

## References

1. Ashton, R. A., Joseph, V., Hoogen, L. L. van den, Tetteh, K. K. A., Stresman, G., Worges, M., Druetz, T., Chang, M. A., Rogier, E., Lemoine, J. F., Drakeley, C., & Eisele, T. P. (2020). Risk Factors for Malaria Infection and Seropositivity in the Elimination Area of Grand’Anse, Haiti: A Case–Control Study among Febrile Individuals Seeking Treatment at Public Health Facilities. The American Journal of Tropical Medicine and Hygiene, 103(2), 767–777. 10.4269/ajtmh.20-0097

2. Druetz, T., Stresman, G., Ashton, R. A., van den Hoogen, L. L., Joseph, V., Fayette, C., Monestime, F., Hamre, K. E., Chang, M. A., Lemoine, J. F., Drakeley, C., & Eisele, T. P. (2020). Programmatic options for monitoring malaria in elimination settings: Easy access group surveys to investigate *Plasmodium falciparum* epidemiology in two regions with differing endemicity in Haiti. BMC Medicine, 18(1), 141. 10.1186/s12916-020-01611-z

3. Bardosh, K., Desir, L., Jean, L., Yoss, S., Poovey, B., Nute, A., de Rochars, M. V. B., Telfort, M.-A., Benoit, F., Chery, G., Charlotin, M. C., & Noland, G. S. (2023). Evaluating a community engagement model for malaria elimination in Haiti: Lessons from the community health council project (2019–2021). Malaria Journal, 22(1), 47. 10.1186/s12936-023-04471-z

4. Moise, K., Achille, A. M., Duvilaire, R., Fedna, J., Thermidor, R., Bourdeau, B., Nestor, J. M., Lerebours, G., Henrys, J. H., & Raccurt, C. (2024). Access and coverage of malaria services in Haiti in the context of elimination: A scoping review of the literature. BMC Health Services Research, 24(1), 1588. 10.1186/s12913-024-12063-z

5. Jacobs, L. D., Judd, T. M., & Bhutta, Z. A. (2016). Addressing the Child and Maternal Mortality Crisis in Haiti through a Central Referral Hospital Providing Countrywide Care. The Permanente Journal, 20(2), 59–70. 10.7812/TPP/15-116

6. Boncy, P. J., Adrien, P., Lemoine, J. F., Existe, A., Henry, P. J., Raccurt, C., Brasseur, P., Fenelon, N., Dame, J. B., Okech, B. A., Kaljee, L., Baxa, D., Prieur, E., El Badry, M. A., Tagliamonte, M. S., Mulligan, C. J., Carter, T. E., Beau de Rochars, V. M., Lutz, C., … Zervos, M. J. (2015). Malaria elimination in Haiti by the year 2020: An achievable goal? Malaria Journal, 14(1), 237. 10.1186/s12936-015-0753-9

7. Weppelmann, T. A., von Fricken, M. E., Lam, B., Telisma, T., Existe, A., Lemoine, J. F., Larkin, J., & Okech, B. A. (2016). Sparse serological evidence of *Plasmodium vivax* transmission in the Ouest and Sud-Est departments of Haiti. Acta Tropica, 162, 27–34. 10.1016/j.actatropica.2016.05.011

8. Lindo, J. F., Bryce, J. H., Ducasse, M. B., Howitt, C., Barrett, D. M., Morales, J. L., Ord, R., Burke, M., Chiodini, P. L., & Sutherland, C. J. (2007). *Plasmodium malariae* in Haitian Refugees, Jamaica. Emerging Infectious Diseases, 13(6), 931. 10.3201/eid1306.061227

9. van den Hoogen, L. L., Herman, C., Présumé, J., Romilus, I., Existe, A., Boncy, J., Joseph, V., Stresman, G., Tetteh, K. K. A., Drakeley, C., Chang, M. A., Lemoine, J. F., Eisele, T. P., Rogier, E., & Ashton, R. A. (2021). Rapid Screening for Non-falciparum Malaria in Elimination Settings Using Multiplex Antigen and Antibody Detection: Post Hoc Identification of *Plasmodium malariae* in an Infant in Haiti. The American Journal of Tropical Medicine and Hygiene, 104(6), 2139–2145. 10.4269/ajtmh.20-1450

10. Bardach, A., Ciapponi, A., Rey-Ares, L., Rojas, J. I., Mazzoni, A., Glujovsky, D., Valanzasca, P., Romano, M., Elorriaga, N., Juri, M. J. D., & Boulos, M. (2015). Epidemiology of Malaria in Latin America and the Caribbean from 1990 to 2009: Systematic Review and Meta-Analysis. Value in Health Regional Issues, 8, 69–79. 10.1016/j.vhri.2015.05.002

11. Hay, S. I., & Snow, R. W. (2006). The Malaria Atlas Project: Developing Global Maps of Malaria Risk. PLOS Medicine, 3(12), e473. 10.1371/journal.pmed.0030473

12. Bento, I., Parrington, B. A., Pascual, R., Goldberg, A. S., Wang, E., Liu, H., Borrmann, H., Zelle, M., Coburn, N., Takahashi, J. S., Elias, J. E., Mota, M. M., & Rijo-Ferreira, F. (2025). Parasite and vector circadian clocks mediate efficient malaria transmission. Nature Microbiology, 10(4), 882–896. 10.1038/s41564-025-01949-1

13. World Malaria Report 2024 (WHO, 2024).

14. Ribeiro, J. M. C., & Francischetti, I. M. B. (2003). Role of Arthropod Saliva in Blood Feeding: Sialome and Post-Sialome Perspectives. Annual Reviews in Entomology, 48, 73–88. 10.1146/annurev.ento.48.060402.102812

15. Londono-Renteria, B. L., Eisele, T. P., Keating, J., James, M. A., & Wesson, D. M. (2010). Antibody response against *Anopheles albimanus* (Diptera: Culicidae) salivary protein as a measure of mosquito bite exposure in Haiti. Journal of Medical Entomology, 47(6), 1156–1163. 10.1603/me09240

16. Londono-Renteria, B., Cardenas, J.C., Cardenas, L.D., Christofferson, R.C., Chisenhall, D.M., Wesson, D.M., McCracken, M.K., Carvajal, D., & Mores, C.N. (2013). Use of Anti-*Aedes aegypti* Salivary Extract Antibody Concentration to Correlate Risk of Vector Exposure and Dengue Transmission Risk in Colombia. PLoS ONE, 8, e81211. 10.1371/journal.pone.0081211.

17. Frederick, J., Saint Jean, Y., Lemoine, J. F., Dotson, E. M., Mace, K. E., Chang, M., Slutsker, L., Le Menach, A., Beier, J. C., Eisele, T. P., Okech, B. A., Beau de Rochars, V. M., Carter, K. H., Keating, J., & Impoinvil, D. E. (2016). Malaria vector research and control in Haiti: A systematic review. Malaria Journal, 15, 376. 10.1186/s12936-016-1436-x

18. Joseph, V., Sutcliffe, A., Leite, L., Czeher, C., Druetz, T., Rogier, E., Eisele, T. P., Lemoine, J. F., Chang, M., Impoinvil, D., & Ashton, R. A. (2025). Entomological Profiles of Households in *Plasmodium falciparum* Case Foci and Comparison Areas in Grand’Anse, Haiti. The American Journal of Tropical Medicine and Hygiene, tpmd240478. 10.4269/ajtmh.24-0478

19. Desenfant P., Molez J.-F., Richard A., Jacques J.-R., Magloire R., & Duverseau Y.T. (1987). Le paludisme en Haïti. 1. Sites d’étude et mise en évidence de sporozoïtes chez *Anopheles albimanus* Wiedmann, 1820. Cah Orstom Sér Ent Méd Parasitol, 25, 69–73.

20. Molez J.-F., Desenfant P., Pajot F.-X., Jacques J.-R., Duverseau Y., & Saint-Jean Y. (1987). Le paludisme en Haïti. 2. Présence d’Anopheles (A.) pseudopunctipennis Theobald, 1901. Première mise en évidence sur l’île d’Hispaniola. Cah Orstom Sér Ent Méd Parasitol, 25, 75–81.

21. Sabrosky C.W., McDaniel G. E., Reider R.F. (1946). A high rate of natural Plasmodium infection in *Anopheles crucians*. Science, 104(2698), 247. https://www.ncbi.nlm.nih.gov/pubmed/21065118

22. González, C., Molina, A. G., León, C., Salcedo, N., Rondón, S., Paz, A., Atencia, M. C., Tovar, C., & Ortiz, M. (2017). Entomological characterization of malaria in northern Colombia through vector and parasite species identification, and analyses of spatial distribution and infection rates. Malaria Journal, 16, 431. 10.1186/s12936-017-2076-5

23. Loyola, E. G., Arredondo, J. I., Rodríguez, M. H., Brown, D. N., & Vaca-Marin, M. A. (1991). *Anopheles vestitipennis*, the probable vector of *Plasmodium vivax* in the Lacandon forest of Chiapas, México. Transactions of The Royal Society of Tropical Medicine and Hygiene, 85(2), 171–174. 10.1016/0035-9203(91)90010-V

24. Fernandez-Salas, I., Rodriguez, M. H., Roberts, D. R., Rodriguez, M. C., & Wirtz, R. A. (1994). Bionomics of adult *Anopheles pseudopunctipennis* (Diptera: Culicidae) in the Tapachula foothills area of southern Mexico. Journal of Medical Entomology, 31(5), 663–670. 10.1093/jmedent/31.5.663

25. DeVita, T. N., Morrison, A. M., Stanek, D., Drennon, M., Sarney, E., Brennan, W., Tomson, K., Blackmore, C., Broussard, K., Duwell, M., Blythe, D., Rothfeldt, L., Dulski, T., Blount, K., Ledford, S., Blackburn, D., Wallender, E., Barratt, J. L. N., Raphael, B. H., … McElroy, P. D. (2025). Public Health Response to the First Locally Acquired Malaria Outbreaks in the US in 20 Years. JAMA Network Open, 8(10), e2535719. 10.1001/jamanetworkopen.2025.35719

26. Grieco, J. P., Achee, N. L., Andre, R. G., & Roberts, D. R. (2002). Host feeding preferences of *Anopheles* species collected by manual aspiration, mechanical aspiration, and from a vehicle-mounted trap in the Toledo District, Belize, Central America. Journal of the American Mosquito Control Association, 18(4), 307–315.

27. Earle W.C. (1936). *Anopheles grabhamii* (Theobald), a possible vector of malaria. Bol. Assoc. Med. P.R, 28, 228–232.

28. Lardeux, F., Loayza, P., Bouchité, B., & Chavez, T. (2007). Host choice and human blood index of *Anopheles pseudopunctipennis* in a village of the Andean valleys of Bolivia. Malaria Journal, 6(1), 8. 10.1186/1475-2875-6-8

29. Loyola, E. G., González-Cerón, L., Rodríguez, M. H., Arredondo-Jiménez, J. I., Bennett, S., & Bown, D. N. (1993). *Anopheles albimanus* (Diptera: Culicidae) Host Selection Patterns in Three Ecological Areas of the Coastal Plains of Chiapas, Southern Mexico. Journal of Medical Entomology, 30(3), 518–523. 10.1093/jmedent/30.3.518

30. Arredondo-Jimenez, J. I., Bown, D. N., Rodriguez, M. H., Villarreal, C., Loyola, E. G., & Frederickson, C. E. (1992). Tests for the existence of genetic determination or conditioning in host selection by *Anopheles albimanus* (Diptera: Culicidae). Journal of Medical Entomology, 29(5), 894–897. 10.1093/jmedent/29.5.894

31. Garrett-Jones, C., Boreham, P. F. L., & Pant, C. P. (1980). Feeding habits of anophelines (Diptera: Culicidae) in 1971–78, with reference to the human blood index: a review. Bulletin of Entomological Research, 70(2), 165–185. 10.1017/S0007485300007422

32. Schneider, B. S., Mathieu, C., Peronet, R., & Mécheri, S. (2011). *Anopheles stephensi* Saliva Enhances Progression of Cerebral Malaria in a Murine Model. Vector-Borne and Zoonotic Diseases, 11(4), 423–432. 10.1089/vbz.2010.0120

33. Donovan, M. J., Messmore, A. S., Scrafford, D. A., Sacks, D. L., Kamhawi, S., & McDowell, M. A. (2007). Uninfected Mosquito Bites Confer Protection against Infection with Malaria Parasites. Infection and Immunity, 75(5), 2523–2530. 10.1128/IAI.01928-06

34. Schneider, B. S., McGee, C. E., Jordan, J. M., Stevenson, H. L., Soong, L., & Higgs, S. (2007). Prior Exposure to Uninfected Mosquitoes Enhances Mortality in Naturally-Transmitted West Nile Virus Infection. PLoS ONE, 2(11), e1171. 10.1371/journal.pone.0001171

35. Vittor, A. Y., Pan, W., Gilman, R. H., Tielsch, J., Glass, G., Shields, T., Sánchez-Lozano, W., Pinedo, V. V., Salas-Cobos, E., Flores, S., & Patz, J. A. (2009). Linking Deforestation to Malaria in the Amazon: Characterization of the Breeding Habitat of the Principal Malaria Vector, *Anopheles darlingi*. The American Journal of Tropical Medicine and Hygiene, 81(1), 5–12.

36. Lindsay, S., Ansell, J., Selman, C., Cox, V., Hamilton, K., & Walraven, G. (2000). Effect of pregnancy on exposure to malaria mosquitoes. The Lancet, 355(9219), 1972. 10.1016/S0140-6736(00)02334-5

37. Lacroix, R., Mukabana, W. R., Gouagna, L. C., & Koella, J. C. (2005). Malaria Infection Increases Attractiveness of Humans to Mosquitoes. PLOS Biology, 3(9), e298. 10.1371/journal.pbio.0030298

38. Kwi, P. N., Ewane, E. E., Moyeh, M. N., Tangi, L. N., Ntui, V. N., Zeukeng, F., Sofeu-Feugaing, D. D., Achidi, E. A., Cho-Ngwa, F., Amambua-Ngwa, A., Bigoga, J. D., & Apinjoh, T. O. (2022). Diversity and behavioral activity of Anopheles mosquitoes on the slopes of Mount Cameroon. Parasites & Vectors, 15(1), 344. 10.1186/s13071-022-05472-8

39. Sallum, M. A. M., Obando, R. G., Carrejo, N., & Wilkerson, R. C. (2020). Identification keys to the *Anopheles* mosquitoes of South America (Diptera: Culicidae). I. Introduction. Parasites & Vectors, 13, 583. 10.1186/s13071-020-04298-6

40. Londono-Renteria, B., Montiel, J., Calvo, E., Tobón-Castaño, A., Valdivia, H. O., Escobedo-Vargas, K., Romero, L., Bosantes, M., Fisher, M. L., Conway, M. J., Vásquez, G. M., & Lenhart, A. E. (2020). Antibody Responses Against *Anopheles darlingi* Immunogenic Peptides in *Plasmodium* Infected Humans. Frontiers in Cellular and Infection Microbiology, 10. 10.3389/fcimb.2020.00455

41. Londono-Renteria, B., Drame, P. M., Montiel, J., Vasquez, A. M., Tobón-Castaño, A., Taylor, M., Vizcaino, L., & Lenhart, A. E. (2020). Identification and Pilot Evaluation of Salivary Peptides from *Anopheles albimanus* as Biomarkers for Bite Exposure and Malaria Infection in Colombia. International Journal of Molecular Sciences, 21(3), Article 3. 10.3390/ijms21030691

42. Poinsignon, A., Cornelie, S., Ba, F., Boulanger, D., Sow, C., Rossignol, M., Sokhna, C., Cisse, B., Simondon, F., & Remoue, F. (2009). Human IgG response to a salivary peptide, gSG6-P1, as a new immuno-epidemiological tool for evaluating low-level exposure to *Anopheles bites*. Malaria Journal, 8(1), 198. 10.1186/1475-2875-8-198

43. Poinsignon, A., Cornelie, S., Mestres-Simon, M., Lanfrancotti, A., Rossignol, M., Boulanger, D., Cisse, B., Sokhna, C., Arcà, B., Simondon, F., & Remoue, F. (2008). Novel Peptide Marker Corresponding to Salivary Protein gSG6 Potentially Identifies Exposure to *Anopheles* Bites. PLoS ONE, 3(6), e2472. 10.1371/journal.pone.0002472

43. Londono-Renteria, B., Drame, P. M., Weitzel, T., Rosas, R., Gripping, C., Cardenas, J. C., Alvares, M., Wesson, D. M., Poinsignon, A., Remoue, F., & Colpitts, T. M. (2015). *An. gambiae* gSG6-P1 evaluation as a proxy for human-vector contact in the Americas: A pilot study. Parasites & Vectors, 8, 533. 10.1186/s13071-015-1160-3

45. Calvo, E., Pham, V. M., Marinotti, O., Andersen, J. F., & Ribeiro, J. M. (2009). The salivary gland transcriptome of the neotropical malaria vector *Anopheles darlingi* reveals accelerated evolution of genes relevant to hematophagy. BMC Genomics, 10, 57. 10.1186/1471-2164-10-57

46. Calvo, E., Andersen, J., Francischetti, I. M., De L. Capurro, M., DeBianchi, A. G., James, A. A., Ribeiro, J. M. C., & Marinotti, O. (2004). The transcriptome of adult female *Anopheles darlingi* salivary glands. Insect Molecular Biology, 13(1), 73–88. 10.1111/j.1365-2583.2004.00463.x

47. Arcà, B., Lombardo, F., Struchiner, C. J., & Ribeiro, J. M. C. (2017). Anopheline salivary protein genes and gene families: An evolutionary overview after the whole genome sequence of sixteen *Anopheles* species. BMC Genomics, 18, 153. 10.1186/s12864-017-3579-8

48. Rizzo, C., Ronca, R., Fiorentino, G., Mangano, V. D., Sirima, S. B., Nèbiè, I., Petrarca, V., Modiano, D., & Arcà, B. (2011). Wide cross-reactivity between *Anopheles gambiae* and *Anopheles funestus* SG6 salivary proteins supports exploitation of gSG6 as a marker of human exposure to major malaria vectors in tropical Africa. Malaria Journal, 10(1), 206. 10.1186/1475-2875-10-206

49. Rizzo, C., Ronca, R., Fiorentino, G., Verra, F., Mangano, V., Poinsignon, A., Sirima, S. B., Nèbiè, I., Lombardo, F., Remoue, F., Coluzzi, M., Petrarca, V., Modiano, D., & Arcà, B. (2011). Humoral Response to the *Anopheles gambiae* Salivary Protein gSG6: A Serological Indicator of Exposure to Afrotropical Malaria Vectors. PLoS ONE, 6(3), e17980. 10.1371/journal.pone.0017980

50. van den Hoogen, L. L., Stresman, G., Présumé, J., Romilus, I., Mondélus, G., Elismé, T., Existe, A., Hamre, K. E. S., Ashton, R. A., Druetz, T., Joseph, V., Beeson, J. G., Singh, S. K., Boncy, J., Eisele, T. P., Chang, M. A., Lemoine, J. F., Tetteh, K. K. A., Rogier, E., & Drakeley, C. (2020). Selection of Antibody Responses Associated With *Plasmodium falciparum* Infections in the Context of Malaria Elimination. Frontiers in Immunology, 11, 928. 10.3389/fimmu.2020.00928

51. van den Hoogen, L. L., Walk, J., Oulton, T., Reuling, I. J., Reiling, L., Beeson, J. G., Coppel, R. L., Singh, S. K., Draper, S. J., Bousema, T., Drakeley, C., Sauerwein, R., & Tetteh, K. K. A. (2019). Antibody Responses to Antigenic Targets of Recent Exposure Are Associated With Low-Density Parasitemia in Controlled Human *Plasmodium falciparum* Infections. Frontiers in Microbiology, 9, 3300. 10.3389/fmicb.2018.03300

52. Greenhouse, B., Smith, D. L., Rodríguez-Barraquer, I., Mueller, I., & Drakeley, C. J. (2018). Taking Sharper Pictures of Malaria with CAMERAs: Combined Antibodies to Measure Exposure Recency Assays. The American Journal of Tropical Medicine and Hygiene, 99(5), 1120–1127. 10.4269/ajtmh.18-0303

53. Lee, S.-K., Han, J.-H., Park, J.-H., Ha, K.-S., Park, W. S., Hong, S.-H., Na, S., Cheng, Y., & Han, E.-T. (2019). Evaluation of antibody responses to the early transcribed membrane protein family in *Plasmodium vivax*. Parasites & Vectors, 12(1), 594. 10.1186/s13071-019-3846-4

54. Druetz, T., van den Hoogen, L., Stresman, G., Joseph, V., Hamre, K. E. S., Fayette, C., Monestime, F., Presume, J., Romilus, I., Mondélus, G., Elismé, T., Cooper, S., Impoinvil, D., Ashton, R. A., Rogier, E., Existe, A., Boncy, J., Chang, M. A., Lemoine, J. F., … Eisele, T. P. (2022). Etramp5 as a useful serological marker in children to assess the immediate effects of mass drug campaigns for malaria. BMC Infectious Diseases, 22, 643. 10.1186/s12879-022-07616-8

55. Dismer, A. M., Lemoine, J. F., Jean Baptiste, M., Mace, K. E., Impoinvil, D. E., Vanden Eng, J., & Chang, M. A. (2021). Detecting Malaria Hotspots in Haiti, a Low-Transmission Setting. The American Journal of Tropical Medicine and Hygiene, 104(6), 2108–2116. 10.4269/ajtmh.20-0465

56. Rogier, E., van den Hoogen, L., Herman, C., Gurrala, K., Joseph, V., Stresman, G., Presume, J., Romilus, I., Mondelus, G., Elisme, T., Ashton, R., Chang, M., Lemoine, J. F., Druetz, T., Eisele, T. P., Existe, A., Boncy, J., Drakeley, C., & Udhayakumar, V. (2019). High-throughput malaria serosurveillance using a one-step multiplex bead assay. Malaria Journal, 18, 402. 10.1186/s12936-019-3027-0

57. Rogier, E., Moss, D. M., Chard, A. N., Trinies, V., Doumbia, S., Freeman, M. C., & Lammie, P. J. (2017). Evaluation of Immunoglobulin G Responses to Plasmodium falciparum and Plasmodium vivax in Malian School Children Using Multiplex Bead Assay. The American Journal of Tropical Medicine and Hygiene, 96(2), 312–318. 10.4269/ajtmh.16-0476

58. R Core Team (2024). R: A Language and Environment for Statistical Computing. R Foundation for Statistical Computing, Vienna, Austria. <https://www.R-project.org/>.

59. Pebesma E, Bivand R (2023). Spatial Data Science With Applications in R. Chapman & Hall. https://r-spatial.org/book/.

61. Gräler B, Pebesma E, Heuvelink G (2016). Spatio-Temporal Interpolation using gstat. The R Journal, 8, 204–218. https://journal.r-project.org/archive/2016/RJ-2016-014/index.html.

62. Samson, D. M., Archer, R. S., Alimi, T. O., Arheart, K. K., Impoinvil, D. E., Oscar, R., Fuller, D. O., & Qualls, W. A. (2015). New baseline environmental assessment of mosquito ecology in northern Haiti during increased urbanization. Journal of Vector Ecology: Journal of the Society for Vector Ecology, 40(1), 46–58. 10.1111/jvec.12131

63. Lombardo, F., Ronca, R., Rizzo, C., Mestres-Simòn, M., Lanfrancotti, A., Currà, C., Fiorentino, G., Bourgouin, C., Ribeiro, J. M. C., Petrarca, V., Ponzi, M., Coluzzi, M., & Arcà, B. (2009). The *Anopheles gambiae* salivary protein gSG6: An anopheline-specific protein with a blood-feeding role. Insect Biochemistry and Molecular Biology, 39(7), 457–466. 10.1016/j.ibmb.2009.04.006

64. Montiel, J., Carbal, L. F., Tobón-Castaño, A., Vásquez, G. M., Fisher, M. L., & Londono-Rentería, B. (2020). IgG antibody response against *Anopheles* salivary gland proteins in asymptomatic *Plasmodium* infections in Narino, Colombia. Malaria Journal, 19(1), 42. 10.1186/s12936-020-3128-9

65. Fontaine, A., Fusaï, T., Briolant, S., Buffet, S., Villard, C., Baudelet, E., Pophillat, M., Granjeaud, S., Rogier, C., & Almeras, L. (2012). *Anopheles* salivary gland proteomes from major malaria vectors. BMC Genomics, 13, 614. 10.1186/1471-2164-13-614

66. Doolan, D. L., Dobaño, C., & Baird, J. K. (2009). Acquired Immunity to Malaria. Clinical Microbiology Reviews, 22(1), 13–36. 10.1128/CMR.00025-08

67. Baird, J. K., Purnomo, Basri, H., Bangs, M. J., Andersen, E. M., Jones, T. R., Masbar, S., Harjosuwarno, S., Subianto, B., & Arbani, P. R. (1993). Age-Specific Prevalence of *Plasmodium falciparum* Among Six Populations with Limited Histories of Exposure to Endemic Malaria. The American Journal of Tropical Medicine and Hygiene, 49(6), 707–719. 10.4269/ajtmh.1993.49.707

68. Asare, K. K., Kwapong, S. S., Tey, P., Sackey, V., Nuvor, S. V., & Amoah, L. E. (2024). *Plasmodium Falciparum* and mosquito vector IgG patterns across suspected malaria cases in Ghana. BMC Infectious Diseases, 24, 1374. 10.1186/s12879-024-10248-9

68. Jaramillo-Underwood, A., Impoinvil, D., Sutcliff, A., Hamre, K. E. S., Joseph, V., van den Hoogen, L., Lemoine, J. F., Ashton, R. A., Chang, M. A., Existe, A., Boncy, J., Drakeley, C., Stresman, G., Druetz, T., Eisele, T., & Rogier, E. (2022). Factors Associated With Human IgG Antibody Response to Anopheles albimanus Salivary Gland Extract, Artibonite Department, Haiti, 2017. The Journal of Infectious Diseases, 226(8), 1461–1469. 10.1093/infdis/jiac245

70. Willen, L., Milton, P., Hamley, J. I. D., Walker, M., Osei-Atweneboana, M. Y., Volf, P., Basáñez, M.-G., & Courtenay, O. (2022). Demographic patterns of human antibody levels to *Simulium damnosum s.l.* saliva in onchocerciasis-endemic areas: An indicator of exposure to vector bites. PLOS Neglected Tropical Diseases, 16(1), e0010108. 10.1371/journal.pntd.0010108

71. Jaramillo-Underwood, A., Herman, C., Impoinvil, D., Sutcliff, A., Knipes, A., Worrell, C. M., Fox, L. M., Desir, L., Fayette, C., Javel, A., Monestime, F., Mace, K. E., Chang, M. A., Lemoine, J. F., Won, K., Udhayakumar, V., & Rogier, E. (2022). Spatial, environmental, and individual associations with *Anopheles albimanus* salivary antigen IgG in Haitian children. Frontiers in Cellular and Infection Microbiology, 12, 1033917. 10.3389/fcimb.2022.1033917

72. Sharma, R. K., Singh, M. P., Saha, K. B., Bharti, P. K., Jain, V., Singh, P. P., Silawat, N., Patel, R., Hussain, M., Chand, S. K., Pandey, A., & Singh, N. (2015). Socio-economic & household risk factors of malaria in tribal areas of Madhya Pradesh, central India. The Indian Journal of Medical Research, 141(5), 567–575. 10.4103/0971-5916.159515

73. Mohan, I., Kodali, N. K., Chellappan, S., Karuppusamy, B., Behera, S. K., Natarajan, G., & Balabaskaran Nina, P. (2021). Socio-economic and household determinants of malaria in adults aged 45 and above: Analysis of longitudinal ageing survey in India, 2017–2018. *Malaria Journal*, 20, 306. 10.1186/s12936-021-03840-w

74. Essendi, W. M., Vardo-Zalik, A. M., Lo, E., Machani, M. G., Zhou, G., Githeko, A. K., Yan, G., & Afrane, Y. A. (2019). Epidemiological risk factors for clinical malaria infection in the highlands of Western Kenya. Malaria Journal, 18, 211. 10.1186/s12936-019-2845-4

75. Ricciardi, I. D. (1971). Definicion de los habitos alimentares de anofelinos de Guatemala y Republica Dominicana, por tecnicas de gel-precipitacion. Revista de Microbiologia, 2(3), 107–112.

76. Mekuria, Y., Tidwell, M. A., Williams, D. C., & Mandeville, J. D. (1990). Bionomic studies of the *Anopheles* mosquitoes of Dajabon, Dominican Republic. Journal of the American Mosquito Control Association, 6(4), 651–657.

76. Mburu, M. M., Zembere, K., Mzilahowa, T., Terlouw, A. D., Malenga, T., van den Berg, H., Takken, W., & McCann, R. S. (2021). Impact of cattle on the abundance of indoor and outdoor resting malaria vectors in southern Malawi. Malaria Journal, 20, 353. 10.1186/s12936-021-03885-x

77. Briegel, H., & Hörler, E. (1993). Multiple Blood Meals as a Reproductive Strategy in *Anopheles* (Diptera: Culicidae). Journal of Medical Entomology, 30(6), 975–985. 10.1093/jmedent/30.6.975

78. Integrated Epidemiological Evaluation: Key Findings and Recommendations, Grand’Anse, Haiti 2015-2020. (2021).

